# Assessment of burden and segregation profiles of CNVs in patients with epilepsy

**DOI:** 10.1101/2022.02.17.22271082

**Authors:** Claudia Moreau, Frédérique Tremblay, Stefan Wolking, Alexandre Girard, Catherine Laprise, Fadi F. Hamdan, Jacques L. Michaud, Berge A. Minassian, Patrick Cossette, Simon L. Girard

## Abstract

1

**Objective:** Microdeletions are associated with different forms of epilepsy but show incomplete penetrance, which is not well understood. We aimed to assess whether unmasked variants or double CNVs could explain incomplete penetrance.

**Methods:** We analyzed copy number variants (CNVs) in 603 patients with four different subgroups of epilepsy and 945 controls. CNVs were called from genotypes and validated on whole genome (WGS) or exome sequences (WES). CNV burden difference between patients and controls was obtained by fitting a logistic regression. CNV burden was assessed for small and large (> 1Mb) deletions and duplications and for deletions overlapping different genes set.

**Results:** Large deletions were enriched in genetic generalized epilepsies (GGE) compared to controls. We also found an enrichment of deletions in epilepsy genes and hotspots for GGE. We did not find truncating or functional variants that could have been unmasked by the deletions. We observed a double CNV hit in two patients. One patient also carried a *de novo* deletion in the 22q11.2 hotspot.

**Interpretation:** We could corroborate previous findings of an enrichment of large microdeletions and deletions in epilepsy genes in GGE. We could also replicate that microdeletions show incomplete penetrance. However, we could not validate the hypothesis of unmasked variants nor the hypothesis of double CNVs to explain the incomplete penetrance. We found a *de novo* hit on 22q11.2 that could be of interest. We also observed GGE families carrying a deletion on 15q13.3 hotspot that could be investigated in the Quebec founder population.

## 2 Introduction

Epilepsy has a prevalence of ∼3% and a high socio-economic burden^1^. About half of the affected individuals experience the first seizures during childhood. About 30-40% of epilepsy syndromes are thought to have a genetic background. Yet, monogenic forms of the disease are rare^2–5^ and represent less than 2% of all cases. The larger share of genetic epilepsy syndromes is thought to be polygenic, which has been substantiated by large-scale genetic studies in the past years^6,7^.

Copy number variants (CNVs) are implicated in the etiology of epilepsy, especially in developmental epileptic encephalopathies (DEE) and genetic generalized epilepsies (GGE)^8–19^. These rare CNVs are either occurring at new sites or at genomic hotspots. Most studies on CNVs in epilepsy focused on microdeletions, although microduplications have also been reported in some cases^20,21^. Moreover, except for non-acquired focal epilepsy (NAFE), large CNVs (generally larger than 1 Mb) are significantly enriched in individuals with epilepsy compared to controls^11,22–24^.

The genetic mechanisms by which these CNVs could cause epilepsy or other developmental disorders remain unclear. In the case of microdeletions, several mechanisms have been proposed to explain their incomplete penetrance, including the unmasking of a recessive allele^25^, a non-coding regulatory variant present in the deletion region^26^ or the presence of a second large CNV that could contribute to a more severe phenotype^27^. The advent of whole genome sequencing (WGS) makes it possible to address these hypotheses more systematically.

Here we investigated CNVs as well as deletions in different sets of genes. The burden of CNVs was assessed in individuals with epilepsy, their unaffected family members and population controls using whole-genome genotyping data. The patients and controls mostly derived from the Quebec founder population^28^. This could maximize our odds of identifying events that would be deemed rare or very rare in populations without founder effect^29,30^. Our extensive familial data collection was used to check for segregation and variant dissemination in larger familial clusters. In addition, we validated the identified large microdeletions and analyzed the homologous chromosome for unmasked variants that could explain the reduced penetrance in patients with WGS or whole exome (WES) sequencing data.

## 3 Subjects and Methods

This study was approved by the CHUM research Center (CRCHUM) ethics committee and by the University of Quebec in Chicoutimi ethics board. Written informed consent was obtained from all patients (or their legal guardians for patients under 18) and adult controls.

### 3.1 Phenotyping

The epilepsy cohort was composed of extended families comprising affected and unaffected individuals with GGE or NAFE as well as DEE trios with unaffected parents previously collected in CHUM Research Center and CHU Ste-Justine in Montreal and in the Hospital for Sick Children in Toronto as part of the Canadian Epilepsy Network (CENet) and diagnosed by neurologists. The clinical epilepsy phenotype was classified according to the current classification by the International League against Epilepsy (ILAE)^31^. Detailed phenotyping is reported in Moreau *et al*.^28^. Certain cases were found with an epilepsy phenotype different from the other affected family members (families marked as “mixed”). The unaffected GGE and NAFE family members and DEE trio parents were used as familial controls in addition to French-Canadian controls from the Quebec Reference Sample^32^.

### 3.2 Genotyping

Samples were processed on either the Illumina Omni Express (n.SNVs = 710,000) or the Illumina Omni 2.5 (n.SNVs = 2,500,000 including the Omni Express core). Genotypes of all samples were merged and only positions present on both chips were kept. We further removed SNVs with more than 2% missing sites over all individuals and with HWE p-value < 0.001 using PLINK software^33^ as well as individuals with more than 2% missing SNVs. Individuals with ambiguous sex were removed from the analysis.

### 3.3 CNV calling and filtering and batch correction

A file was generated by the Genome Quebec Innovation Center in Montreal for each genotyped sample including Log-R ratio (LRR) and B allele frequency (BAF) for all SNVs. PennCNV software^34^ was used for CNV calling. Only filtered SNVs were used to generate a custom population B-allele frequency file before calling CNVs. First CNV calling (--qclrrsd 0.3 --qcbafdrift 0.01 --qcwf 0.05) was performed to remove low quality samples, then principal components analysis and batch correction (PC-correction) was applied to LRR as described in Cooper *et al*.^35^ using filtered SNPs outside of telomeric, centromeric and immunoglobulin regions (Supplementary Fig 1). Second, CNV calling was performed on the corrected LRR using --qclrrsd 0.3 --qcbafdrift 0.01 --qcwf 0.05 --numsnp 10 --length 20k --qcnumcnv 50, telomeric, centromeric and immunoglobulin regions were removed and CNVs were merged using default fraction argument of 0.2. Total number of samples, males and females after QC in addition to available WGS and WES are presented in (Table 1). CNVs were also called on 135 complete DEE, GGE, NAFE or mixed trios to look for *de novo* CNV hits.

**Table 1:**
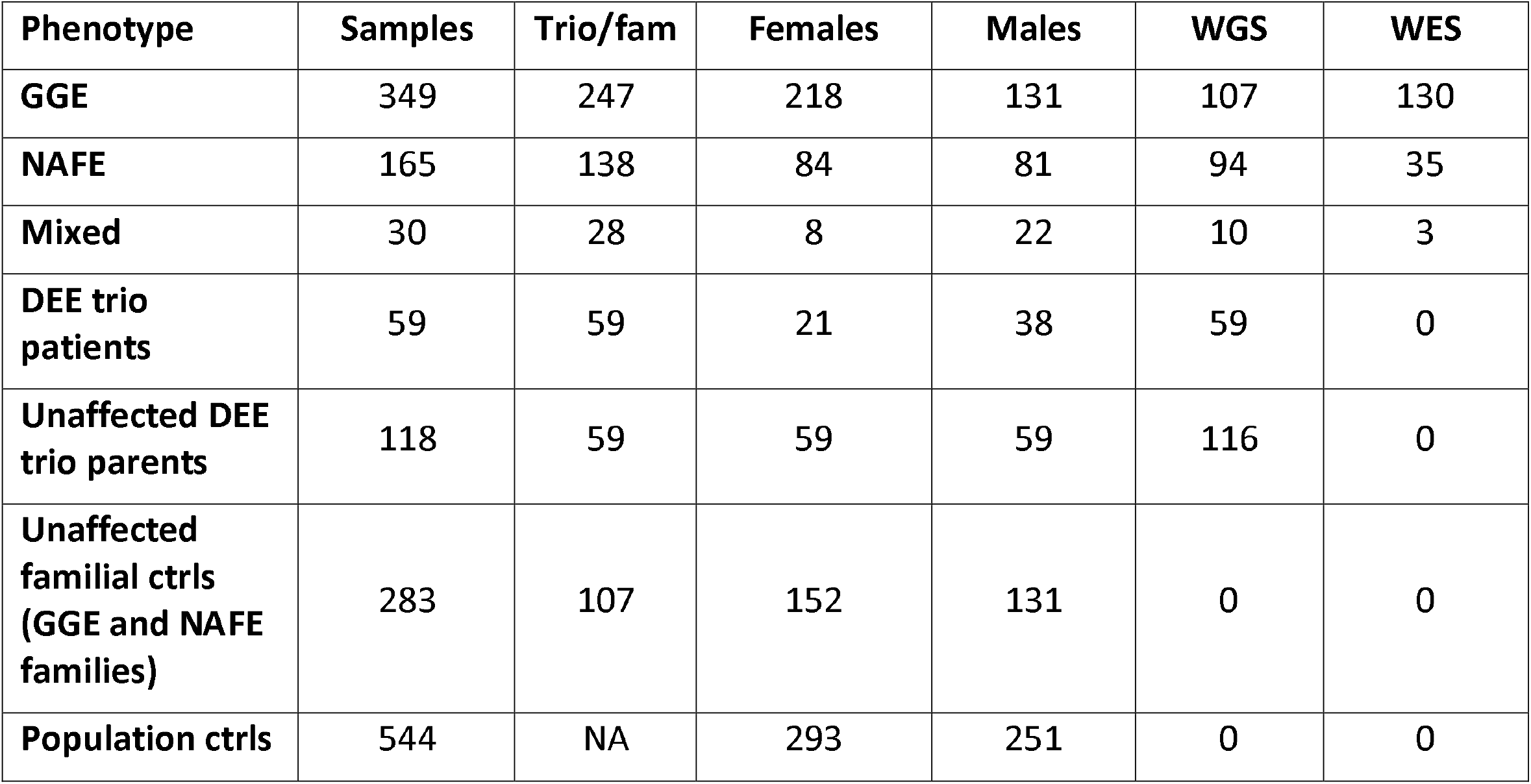
Number of individuals in each group. GGE = genetic generalized epilepsies ; NAFE = non-acquired focal epilepsy ; Mixed = cases with an epilepsy phenotype different from the other affected family members ; DEE = developmental epileptic encephalopathies ; ctrls = controls.

We only considered rare CNVs (<=1%) for further analyses. There were 4,460 such CNVs in our dataset. The CNV frequency was obtained using PLINK^33^ v1.07 –cnv-freqmethod2 0.5 option.

### 3.4 CNV validation

CNVs were validated using either whole genome (WGS) or whole exome (WES) depending on the availability of such sequences and/or segregation in the family. For segregation, CNVs were considered as being the same if they overlapped at least 50%. Duplications and deletions were considered separately. Detailed sequencing methods for WGS and WES are described in Moreau *et al*.^28^ and in Wolking *et al*.^36^ respectively. CNVs on WGS and WES were called using two software, CNVkit^37^ and Control-FREEC^38^. A CNV was considered as validated if called by one of these software and overlap at least 50%. We did not consider a CNV as validated if the length of the WGS or WES call was more than twice the length of the genotyping call to avoid spurious calls.

### 3.5 CNV annotation

PennCNV was used to determine if CNVs were spanning genes (hg19). A CNV was considered to be in the coding region if it overlapped at least 80% of a gene. We also identified 152 genes that were previously associated with epilepsy^39,40^ and 1,804 genes intolerant for protein truncating variants defined as probability of loss-of-function (lof) intolerance (pLI) score > 0.99. Epilepsy hotspots previously identified in epileptic patients^41^ (Supplementary Table 1). A CNV was considered to be in a hotspot if it overlapped at least 50% of a hotspot.

### 3.6 CNV burden

We measured CNV burden for all epilepsy phenotypes for small and large (> 1Mb) rare deletions and duplications separately to evaluate relative contribution on epilepsy type risk. We also looked at rare deletions overlapping genes, epilepsy-associated genes, genes with pLi > 0.99 and known epilepsy hotspots (Supplementary Table 1). To assess for a CNV burden difference between epilepsy cases and controls, we fitted a logistic binomial regression model with sex as covariate using the geekin function of the MESS package (https://cran.r-project.org/web/packages/MESS/index.html) to account for familial relationships. The familial relationships were obtained using PLINK –genome option after pruning. For all burden analyses, odds ratios, 95% confidence intervals (CIs), and significance were calculated by taking the exponential of the logistic regression coefficients. We removed the unaffected DEE parents from the DEE burden analyses. Bonferroni multiple-testing was calculated for 16 tests for both groups of analyses and threshold for significance was 0.003.

### 3.7 Variant calling and annotation

SNVs in microdeletions were called in WGS or WES. We performed joint calling of gvcf files that were merged into a single vcf file using GATK version 3.7□0 (https://gatk.broadinstitute.org/hc/en-us). The vcf file was recalibrated and filtered following the GATK best practice guidelines. SnpEff and SnpSift^42,43^ were used to annotate SNVs. A SNV was considered having a lof or nonsense-mediated mRNA decay (nmd) effect if this effect was seen in more than 90% of the transcripts. All missense variants were considered. We cross-referenced these SNVs in ClinVar (version of June 9^TH^, 2021)^44^ to identify known pathogenic variants. To assess whether non-coding SNVs could have a functional effect, we used ExPecto^45^, a deep learning algorithm that computes the tissue specific effect of variants on gene expression using WGS and WES (although WES are not expected to include many non-coding variants). The computed expression fold change resulting of Expecto analysis was used to identify deleterious variants. We calculated a variation potential directionality score for each gene for three tissues related to epilepsy (amygdala, cortex and hippocampus). Then a constraint violation score was obtained by computing the product of the variation potential directionality score and the predicted expression change for a given SNV. The higher this score is, the more deleterious the SNV is.

## 4 Results

Burden analysis revealed a greater proportion of deletions > 1 Mb in GGE individuals resulting in significant OR (4.97 ; 95% CI 2.5-10.1) against controls (Fig 1). Moreover, we also observed an excess of large deletions compared to large duplications in GGE. No such proportions were observed for duplications. Since microdeletions seem more important in epilepsy^24^, burden analysis was performed only on deletions for different gene sets and epilepsy hotspots (Fig 2). We found an enrichment of deletions in epilepsy genes and hotspots for GGE compared to controls (Supplementary Tables 2 and 3 for detailed CNV description).

**Figure 1:**
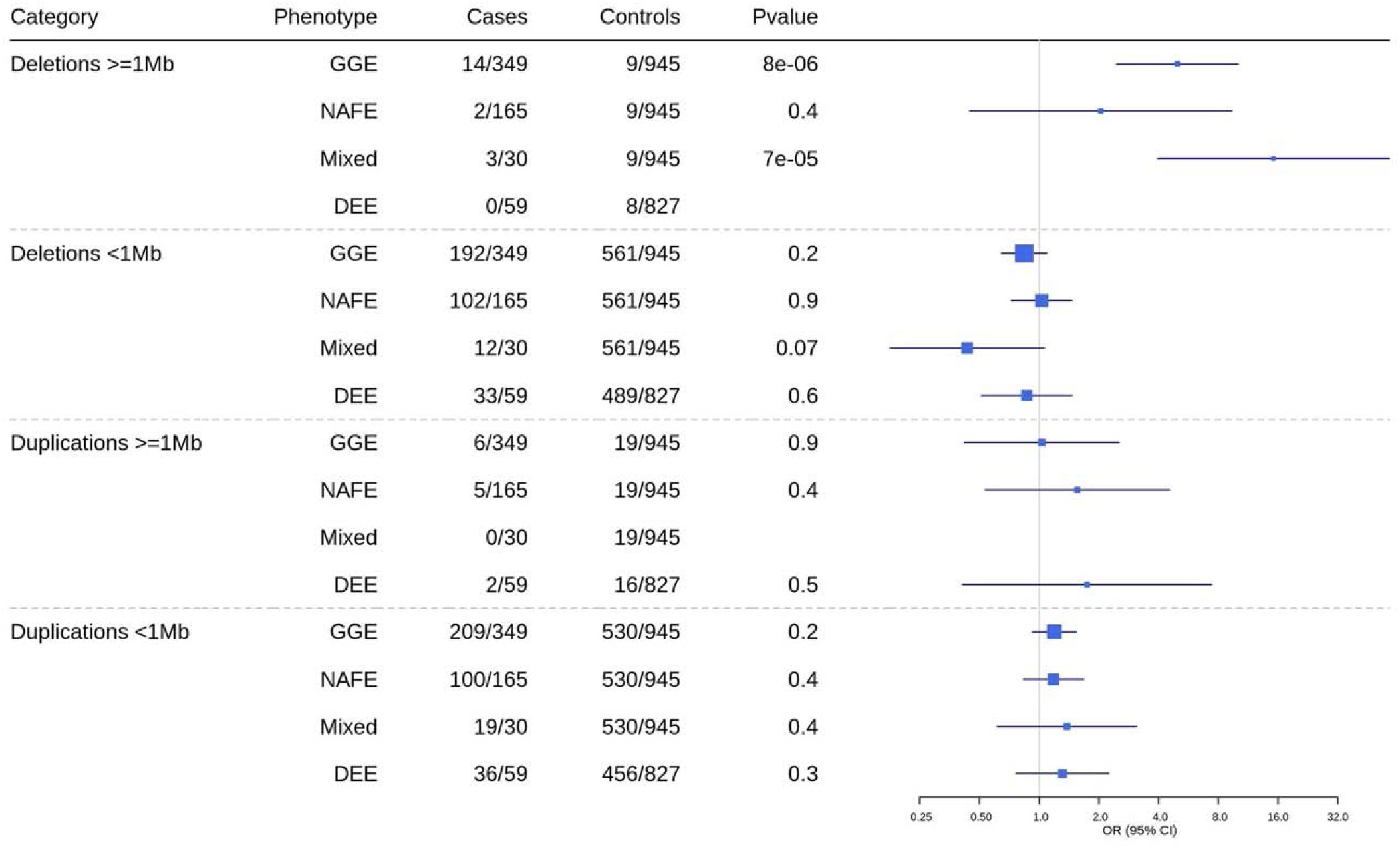
Burden of CNVs by length in epilepsy subgroups. GGE = genetic generalized epilepsies ; NAFE = non-acquired focal epilepsy ; Mixed = cases with an epilepsy phenotype different from the other affected family members ; DEE = developmental epileptic encephalopathies.

**Figure 2:**
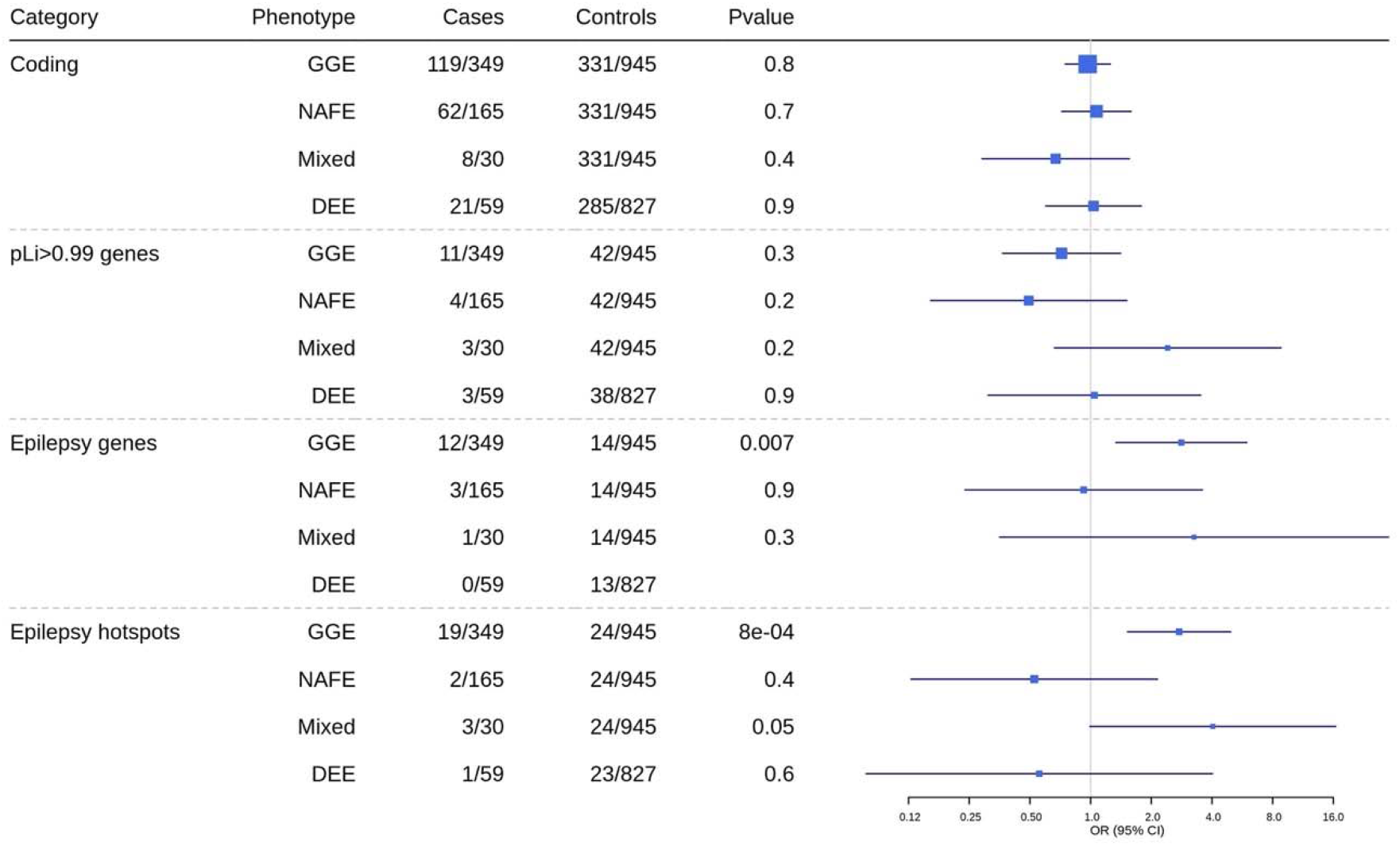
Burden of deletions across different gene sets or hotspots in epilepsy subgroups. GGE = genetic generalized epilepsies ; NAFE = non-acquired focal epilepsy ; Mixed = cases with an epilepsy phenotype different from the other affected family members ; DEE = developmental epileptic encephalopathies.

We further analyzed individuals carrying a large deletion (Table 2 and Supplementary Table 4). All deletions found in patients were located within a gene whereas only 30% of the deletions among the population controls was located in the coding region. Half of the large deletions were found in an epilepsy gene or hotspot (Table 2). All 12 deletions for which we had WGS or WES in addition to genotypes were validated. The remaining deletions were validated by looking at the segregation in the family. Almost all validated deletions for which we had family information were transmitted either by an affected (four transmissions + one plausible transmission) or an unaffected parent (four transmissions) (Supplementary Table 4). Only one *de novo* large deletion could not be validated because the patient did not have WGS data. As reported previously, we document here several cases where known pathological hotspots CNV were either transmitted from an unaffected family member or to a yet unaffected sibling warranting the need to be cautious when using these findings in clinical settings.

**Table 2:** Number of individuals carrying deletions > 1 Mb. Dels = deletions ; pLi = genes intolerant to truncating variants ; GGE = genetic generalized epilepsies ; NAFE = non-acquired focal epilepsy ; Mixed = cases with an epilepsy phenotype different from the other affected family members ; DEE = developmental epileptic encephalopathies ; ctrls = controls.

| Phenotype | Dels<br>><br>1Mb | Coding | pLi<br>><br>0.99 | Epilepsy<br>genes | Epilepsy<br>hotspots | Validated<br>by WGS | Validated<br>by WES | Validated<br>by<br>segregation | Validated<br>overall |
| --- | --- | --- | --- | --- | --- | --- | --- | --- | --- |
| GGE | 14 | 14 | 2 | 7 | 9 | 5 | 4 | 8 | 14 |
| NAFE | 2 | 2 | 1 | 0 | 0 | 0 | 1 | 1 | 2 |
| Mixed | 3 | 3 | 1 | 0 | 1 | 1 | 0 | 1 | 2 |
| DEE trio<br>patients | 0 | 0 | 0 | 0 | 0 | 0 | 0 | 0 | 0 |
| Unaffected<br>DEE trio<br>parents | 1 | 1 | 0 | 0 | 0 | 1 | 0 | 0 | 1 |
| Unaffected<br>familial<br>ctrls (GGE<br>and NAFE<br>families) | 5 | 5 | 2 | 2 | 4 | 0 | 0 | 5 | 5 |
| Population<br>ctrls | 3 | 1 | 0 | 0 | 0 | 0 | 0 | 0 | 0 |

Among individuals carrying a deletion validated by WGS or WES, we looked for variants of interest on the other chromosome that could have been unmasked by the deletion. Missense variants were found (Supplementary Table 5) and were re-validated in IGV^46^. They had to be homozygote, as expected given a deletion on the other chromosome. Most of the missense variants were frequent, with only one variant at less than 1% allele frequency in gnomAD (rs762560584). Moreover, the UNEECON scores^47^, that predict how deleterious a missense variant is, were under 0.15 (not deleterious) for all variants. One non-coding variant (chr15:31195835CAG>C) in a GGE patient had a negative constraint score for the three tested tissues, which implies that it is not likely to be deleterious and is also quite frequent in gnomAD (0.42). The other non-coding variants did not have a constraint score meaning that they are not likely to have any functional effect.

We identified two NAFE patients with a double CNV hit. One had a duplication transmitted by the unaffected mother and a deletion transmitted by the affected father, both on chromosome 7 (chr7:88161734-89838707dup and chr7:108854537-109969407del) and validated by segregation. The second patient had one duplication followed immediately by a 13Mb deletion on chromosome 18 (chr18:63151948-64412293dup and chr18:64525217-77553173del), both validated by WES, but with no family information.

## 5 Discussion

In the present work, we found an excess of deletions of more than 1 Mb in GGE patients compared to controls, and to a lesser extent, in individuals from mixed families, comparable to previous findings^24^. We also found an excess of large deletions compared to duplications in GGE patients, again comparable to previous findings^18,22^. Most of these deletions were located in epilepsy genes or hotspots. Moreover, we found an excess of deletions in epilepsy genes and hotspots in GGE patients which is mostly driven by a deletion on the 15q13.3 recurrent site which is also spanning an epilepsy gene, *CHRNA7* and has been reported previously in GGE patients^8,13^ (OMIM 612001). This deletion in the 15q13.3 hotspot region was present exclusively in seven GGE patients from six different families and two unaffected family members and was not reported in any population control nor in other epilepsy types. It is the only deletion in an epilepsy hotspot that was restricted to patients and their relatives in this study. This could be a variant linked to the founder effect in the Quebec population^48^ and propagated mostly to the affected descendants of a given ancestor. This would need further family and population analyses to validate the transmission scheme of a variant associated to a disease compared to one that is only resulting of the expected transmission in a founder population without any disease association.

The only *de novo* large deletion was found in a patient from a mixed family (DEE in a NAFE family, Supplementary Fig 2). Interestingly, it was found in an epilepsy hotspot, 22q11.2 (OMIM 611867). In addition to DEE, the patient presented a severe intellectual disability and autism like symptoms which are associated with the 22q11.2 deletion. Interestingly, it has been shown that 11% of the 22q11.2 deletion carriers have epilepsy and an additional 59% have seizures or seizure-like symptoms^49^. This could also explain the DEE phenotype within a NAFE family for this patient.

Most of the identified large deletions in epileptic patients were transmitted either by an affected or an unaffected parent, denoting incomplete penetrance^50^ with only one deletion that occurred *de novo*. The validation rate was high in the present study, thanks to the variety of data available for these patients. To test whether the incomplete penetrance of epilepsy-related deletions could be explained by the unmasking of a variant on the other chromosome, we looked at the deletion regions in available WGS and WES for lof and missense variants in addition to variants in ClinVar and variants predicted to affect gene expression using Expecto (see Methods for details). We did not find any evidence of lof or other variants classified as probably pathogenic in ClinVar or affecting gene expression. We found missense variants that are not predicted deleterious according to the annotations in gnomAD and the UNEECON scores^47^ (Supplementary Table 5).

Another hypothesis that has been proposed to explain incomplete penetrance is the double hit hypothesis^50^. Two NAFE patients had a double CNV hits, one patient had both hits on chromosome 7 and the other both on chromosome 18. The former patient’s duplication on chromosome 7 is the most frequent duplication and has been seen in 12 patients and controls in the present dataset. Both CNVs on chromosome 7 affect coding regions, but do not include genes intolerant to truncating variants nor known epilepsy genes or hotspots, so we do not have evidence that these would be associated with the disease. However, the second NAFE patient had both hits, a duplication and a deletion, adjacent on chromosome 18. The deletion was the largest found in our dataset, spanning 13Mb and two genes intolerant to truncating variants, *ZNF236* and *ZNF407* that were associated to chromosome 18q deletion syndrome (OMIM 601808), neurodevelopmental disorders, and intellectual disability^51^, among others. The finding of two CNVs in this case does not necessarily support the double hit hypothesis since it cannot be ruled out that the deletion alone caused the phenotype.

In conclusion we found an excess of large deletions in GGE patients compared to unaffected familial controls from the CENet cohort and population controls from the Quebec Reference Sample, and also compared to the number of duplications in GGE patients. Most of the deletions are located at genomic hotspots in GGE, especially at the 15q13.3 site which could have been brought and disseminate by an ancestor of the Quebec founder population. We also found one *de novo* deletion that could explain the patient’s phenotype and be of interest for the medical follow up. We could not find evidence of deleterious or regulatory variants on the homologous chromosome that would explain the incomplete penetrance of the disease among individuals having large deletions. The double hit hypothesis could not be supported neither although we found two large CNVs in two NAFE patients including one deletion of 13Mb that could be of interest for the patient and the clinician. We found missense variants within the deletion regions that seem not sufficient to explain the disease. Therefore, we think that there might be other genomic or epigenomic causes in addition to large deletions that would explain the incomplete penetrance of epilepsy related microdeletions, although we need more sequencing data to validate these findings.

All authors participated in the study design and reviewed the manuscript. CM, FT and AG performed the analyses. CM wrote the manuscript. SLG supervised the analyses. CL built and manages the SLSJ family cohort which provided the Saguenay population control samples.

## Data Availability

All data produced in the present work are contained in the manuscript

## 6 Acknowledgement

This work was supported by funding from Genome Quebec / Genome Canada as well as from the CIHR (#420021). It was also made possible by Compute Canada resources allocation for the access to storage and computing resources. We are extremely grateful to all patients and their families for participating in this research. We would like to thank Héléne Vézina and Damian Labuda for the Quebec Reference Sample cohort constitution. SW received funding from the German Reserach Foundation (WO-2385/2-1). CL is the director of the Centre intersectoriel en santé durable de l’UQAC (CISD; http://www.uqac.ca/santedurable) and the chairholder of the Canada Research Chair in the Environment and Genetics of Respiratory Diseases and Allergy (http://www.chairs.gc.ca).

## 9 Legends

**Supplementary Figure 1: LRR’s PCA before (left panel) and after (right panel) PC correction**. Symbols represent the different batches and colors the different plates (many plates were sent for genotyping within one batch).

**Supplementary Figure 2: Pedigree of the mixed patient (DEE in a NAFE family) carrying the *de novo* deletion**. Unaffected individuals are in black.

**Supplementary Table 1: Recurrent deletions’ description from Watson *et al***.

**Supplementary Table 2: Deletions in epilepsy genes**.

**Supplementary Table 3: Deletions in epilepsy hotspots**.

**Supplementary Table 4: Description of deletions > 1 Mb**.

**Supplementary Table 5: Unmasked missense variants**.

