## Supplemental Figures for "Assessment of burden and segregation profiles of CNVs in patients with epilepsy"

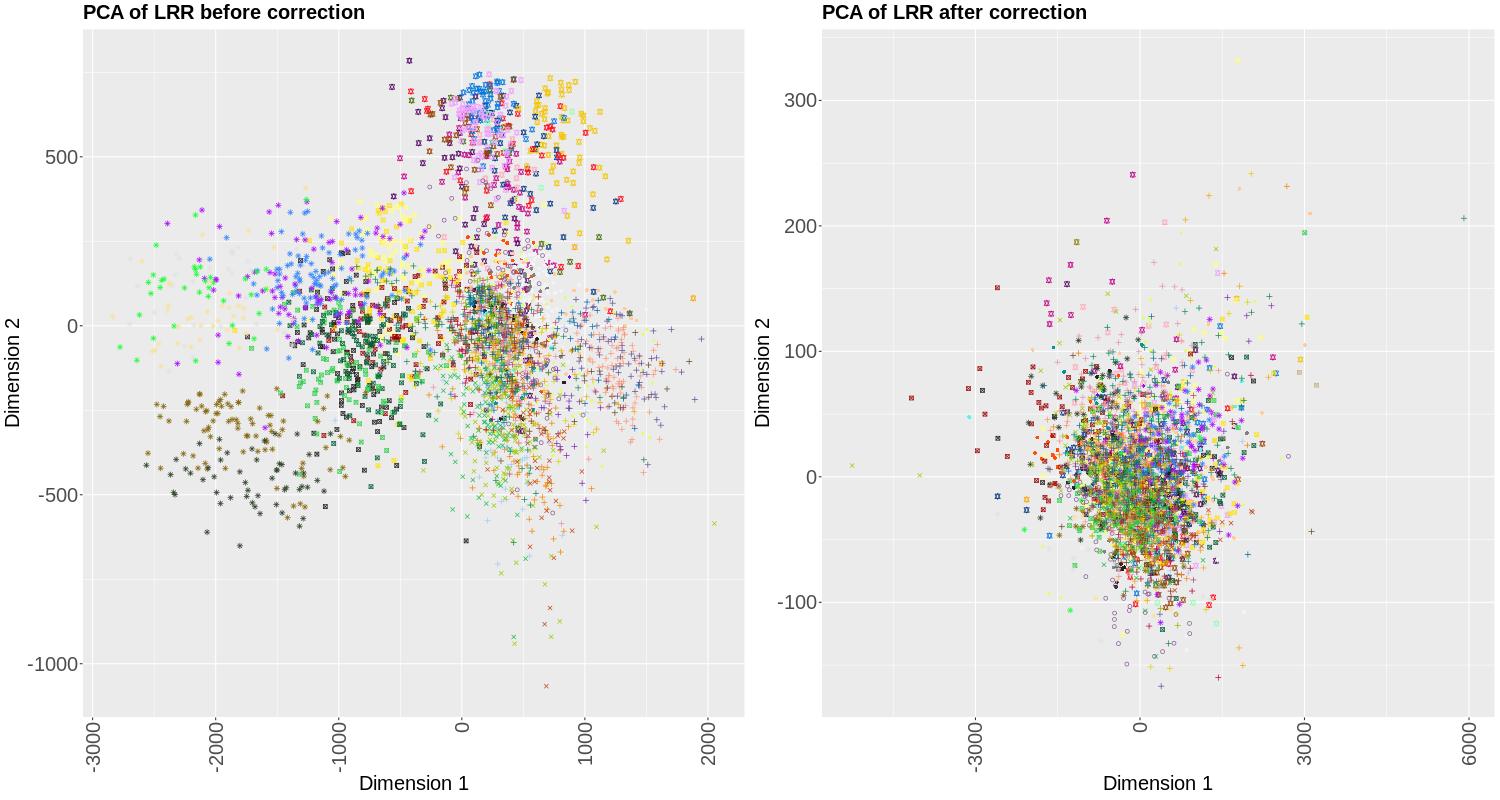

**Supplementary figure 1:** LRR’s PCA before (left panel) and after (right panel) PC correction. Symbols represent the different batches and colors the different plates (many plates were sent for genotyping within one batch).

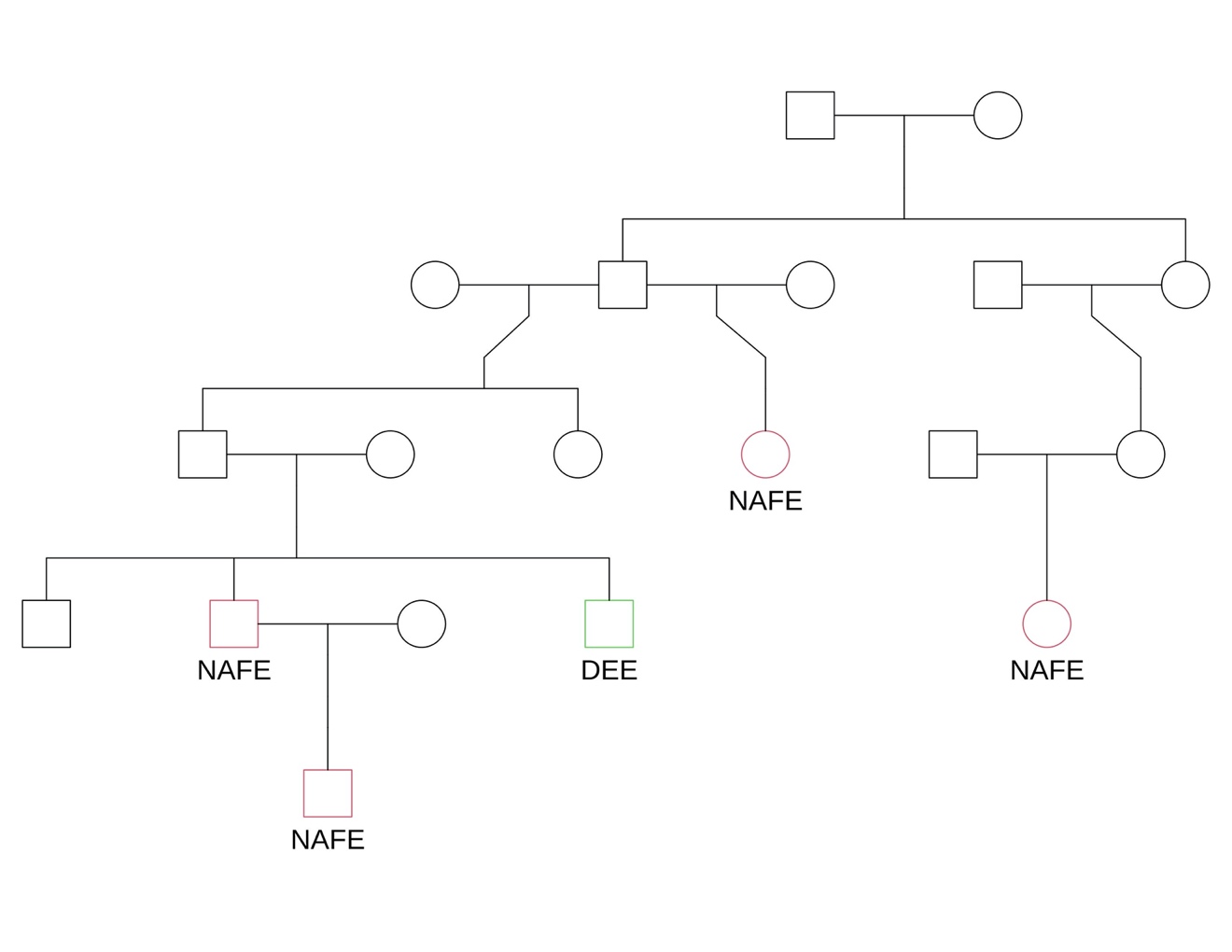

**Supplementary Figure 2:** Pedigree of the mixed patient (DEE in a NAFE family) carrying the *de novo* deletion. Unaffected individuals are in black.
