## Supplemental Table 1 for "Assessment of burden and segregation profiles of CNVs in patients with epilepsy"

| **Chromosomal location** | **Coordinates hg19** | **OMIM** | **Epilepsy/candidate gene** | **Selected reference** |
| --- | --- | --- | --- | --- |
| 1q21.1 | Chr1 :146.5-147.5 |  | *GJA8* | ^1–3^ |
| 2q21.1 | chr2:131.48-131.9 |  |  | ^4^ |
| 7q11.23 distal | chr7:75.07-76.25 | 613729 |  | ^5^ |
| 10q11.21–q11.23 | chr10:42.5-52.5 |  |  | ^6^ |
| 15q11.2 | chr15:21-25.5 | 615656 | *NIPA1,CYFIP1* | ^2,7^ |
| 15q11–q13 | chr15:23-28.5 |  |  | ^8^ |
| 15q13.3 | chr15:31.5-33.5 | 612001 | *CHRNA7* | ^9,10^ |
| 16p11.2 | chr16:28.5-34 | 611913 | *PRRT2* | ^11^ |
| 16p13.11 | chr16:14.9-16.6 |  | *NDE1* | ^7,12^ |
| 17q12 | chr17:32-37.5 | 614527 |  | ^13^ |
| 22q11.2 | chr22:18-25.5 | 611867 | *SNAP29* | ^3^ |

**Supplementary Table 1:** Recurrent deletions’ description from Watson *et al.* ^14^
