## Supplemental Table 2 for "Assessment of burden and segregation profiles of CNVs in patients with epilepsy"

| CNV | Length | Genes | Phenotype | Sex | Family | Coding |
|---|---|---|---|---|---|---|
| chr1:44174081-44446651 | 272571 | ARTN,ATP6V0B,B4GALT2,DPH2,IPO13,ST3GAL3,U6 | GGE | 1 | EG231 | 1 |
| chr14:67146763-67486808 | 340046 | GPHN | GGE | 2 | EG233 | 1 |
| chr15:30936285-32514341 | 1578057 | AK093758,ARHGAP11B,CHRNA7,DKFZp434L187,DQ572979,DQ582939,DQ588973,DQ595055,DQ596686,DQ600342,DQ786280,FAN1,HERC2P10,JB175342,KLF13,LOC100288637,LOC283710,MIR211,MTMR10,OTUD7A,TRPM1 | fam_ctrl | 2 | EG001 | 1 |
| chr15:30936285-32514341 | 1578057 | AK093758,ARHGAP11B,CHRNA7,DKFZp434L187,DQ572979,DQ582939,DQ588973,DQ595055,DQ596686,DQ600342,DQ786280,FAN1,HERC2P10,JB175342,KLF13,LOC100288637,LOC283710,MIR211,MTMR10,OTUD7A,TRPM1 | fam_ctrl | 2 | EG001 | 1 |
| chr15:30936285-32514341 | 1578057 | AK093758,ARHGAP11B,CHRNA7,DKFZp434L187,DQ572979,DQ582939,DQ588973,DQ595055,DQ596686,DQ600342,DQ786280,FAN1,HERC2P10,JB175342,KLF13,LOC100288637,LOC283710,MIR211,MTMR10,OTUD7A,TRPM1 | GGE | 1 | EG001 | 1 |
| chr15:30936285-32514341 | 1578057 | AK093758,ARHGAP11B,CHRNA7,DKFZp434L187,DQ572979,DQ582939,DQ588973,DQ595055,DQ596686,DQ600342,DQ786280,FAN1,HERC2P10,JB175342,KLF13,LOC100288637,LOC283710,MIR211,MTMR10,OTUD7A,TRPM1 | GGE | 2 | EG001 | 1 |
| chr15:30936285-32514341 | 1578057 | AK093758,ARHGAP11B,CHRNA7,DKFZp434L187,DQ572979,DQ582939,DQ588973,DQ595055,DQ596686,DQ600342,DQ786280,FAN1,HERC2P10,JB175342,KLF13,LOC100288637,LOC283710,MIR211,MTMR10,OTUD7A,TRPM1 | GGE | 2 | EG087 | 1 |
| chr15:30936285-32514341 | 1578057 | AK093758,ARHGAP11B,CHRNA7,DKFZp434L187,DQ572979,DQ582939,DQ588973,DQ595055,DQ596686,DQ600342,DQ786280,FAN1,HERC2P10,JB175342,KLF13,LOC100288637,LOC283710,MIR211,MTMR10,OTUD7A,TRPM1 | GGE | 1 | EG282 | 1 |
| chr15:30936285-32514341 | 1578057 | AK093758,ARHGAP11B,CHRNA7,DKFZp434L187,DQ572979,DQ582939,DQ588973,DQ595055,DQ596686,DQ600342,DQ786280,FAN1,HERC2P10,JB175342,KLF13,LOC100288637,LOC283710,MIR211,MTMR10,OTUD7A,TRPM1 | GGE | 1 | EG360 | 1 |
| chr15:30936285-32514341 | 1578057 | AK093758,ARHGAP11B,CHRNA7,DKFZp434L187,DQ572979,DQ582939,DQ588973,DQ595055,DQ596686,DQ600342,DQ786280,FAN1,HERC2P10,JB175342,KLF13,LOC100288637,LOC283710,MIR211,MTMR10,OTUD7A,TRPM1 | GGE | 2 | EG328 | 1 |
| chr15:30936285-32514341 | 1578057 | AK093758,ARHGAP11B,CHRNA7,DKFZp434L187,DQ572979,DQ582939,DQ588973,DQ595055,DQ596686,DQ600342,DQ786280,FAN1,HERC2P10,JB175342,KLF13,LOC100288637,LOC283710,MIR211,MTMR10,OTUD7A,TRPM1 | GGE | 1 | EG430 | 1 |
| chr16:29652488-30192359 | 539872 | AB209061,AK097453,AK097472,AK097527,ALDOA,ASPHD1,BC029255,BC041466,BOLA2,C16orf54,C16orf92,CDIPT,CDIPT-AS1,DOC2A,FAM57B,GDPD3,HIRIP3,INO80E,KCTD13,KIF22,MAPK3,MAZ,MVP,PAGR1,PPP4C,PRRT2,QPRT,SEZ6L2,SPN,TAOK2,TBX6,TMEM219,YPEL3,ZG16 | Mixed | 2 | EG067 | 1 |
| chr16:79095848-79121835 | 25988 | WWOX | ctrl | 2 | NA | 1 |
| chr16:79095848-79121835 | 25988 | WWOX | DEE_trio_parent | 1 | Trio64 | 1 |
| chr16:79130574-79171137 | 40564 | WWOX | fam_ctrl | 2 | EG029 | 1 |
| chr16:79130574-79171137 | 40564 | WWOX | GGE | 2 | EG029 | 1 |
| chr16:79130574-79171137 | 40564 | WWOX | GGE | 1 | EG029 | 1 |
| chr2:50918967-51033295 | 114329 | NRXN1 | ctrl | 2 | NA | 1 |
| chr22:32129550-32240460 | 110911 | DEPDC5,PRR14L | fam_ctrl | 2 | EP007 | 1 |
| chr22:32129550-32240460 | 110911 | DEPDC5,PRR14L | NAFE | 2 | EP007 | 1 |
| chr22:32129550-32255802 | 126253 | DEPDC5,PRR14L | fam_ctrl | 1 | EP007 | 1 |
| chr22:33760604-34150984 | 390381 | LARGE,LARGE-AS1,MIR4764,SNORA50 | fam_ctrl | 1 | EP013 | 1 |
| chr22:33760604-34157526 | 396923 | LARGE,LARGE-AS1,MIR4764,SNORA50 | NAFE | 2 | EP013 | 1 |
| chr22:33760604-34172801 | 412198 | LARGE,LARGE-AS1,MIR4764,SNORA50 | ctrl | 2 | NA | 1 |
| chr22:33760604-34172801 | 412198 | LARGE,LARGE-AS1,MIR4764,SNORA50 | fam_ctrl | 2 | EP031 | 1 |
| chr22:34096135-34194745 | 98611 | LARGE,LARGE-AS1,SNORA50 | ctrl | 2 | NA | 1 |
| chr22:34096135-34194745 | 98611 | LARGE,LARGE-AS1,SNORA50 | NAFE | 1 | EP027 | 1 |
| chr22:34096135-34196626 | 100492 | LARGE,LARGE-AS1,SNORA50 | ctrl | 2 | NA | 1 |
| chr7:147011388-147067719 | 56332 | CNTNAP2,MIR548I4 | ctrl | 1 | NA | 1 |
| chr8:17888770-17977230 | 88461 | ASAH1 | GGE | 2 | EG315 | 1 |

Red = epilepsy genes from:


Berkovic SF, Scheffer IE, Petrou S, et al. A roadmap for precision medicine in the epilepsies. The Lancet Neurology 2015;14(12):1219–28.

Coppola A, Cellini E, Stamberger H, et al. Diagnostic implications of genetic copy number variation in epilepsy plus. Epilepsia 2019;60(4):689–706.
