## Supplemental Table 3 for "Assessment of burden and segregation profiles of CNVs in patients with epilepsy"

| CNV | Length | Genes | Phenotype | Sex | Family | Coding |
|---|---|---|---|---|---|---|
| chr15:24112281-24154189 | 41909 | NOT_FOUND | GGE | 1 | EG124 | 0 |
| chr15:24369643-24468460 | 98818 | AK124131,PWRN2 | ctrl | 1 | NA | 1 |
| chr15:24369643-24468460 | 98818 | AK124131,PWRN2 | ctrl | 2 | NA | 1 |
| chr15:24369643-24468460 | 98818 | AK124131,PWRN2 | ctrl | 2 | NA | 1 |
| chr15:24369643-24468460 | 98818 | AK124131,PWRN2 | ctrl | 1 | NA | 1 |
| chr15:24369643-24468460 | 98818 | AK124131,PWRN2 | DEE | 1 | Trio102 | 1 |
| chr15:24369643-24468460 | 98818 | AK124131,PWRN2 | DEE_trio_parent | 1 | Trio102 | 1 |
| chr15:24369643-24468460 | 98818 | AK124131,PWRN2 | fam_ctrl | 1 | EG395 | 1 |
| chr15:24369643-24468460 | 98818 | AK124131,PWRN2 | GGE | 2 | EG212 | 1 |
| chr15:24369643-24468460 | 98818 | AK124131,PWRN2 | GGE | 2 | EG344 | 1 |
| chr15:24369643-24468460 | 98818 | AK124131,PWRN2 | GGE | 1 | EG433 | 1 |
| chr15:24369643-24649604 | 279962 | AK124131,PWRN2 | NAFE | 2 | EP231 | 1 |
| chr15:24376622-24468460 | 91839 | AK124131,PWRN2 | ctrl | 1 | NA | 1 |
| chr15:25962058-26019039 | 56982 | ATP10A | ctrl | 1 | NA | 1 |
| chr15:30936285-32514341 | 1578057 | AK093758,ARHGAP11B,CHRNA7,DKFZp434L187,DQ572979,DQ582939,DQ588973,DQ595055,DQ596686,DQ600342,DQ786280,FAN1,HERC2P10,JB175342,KLF13,LOC100288637,LOC283710,MIR211,MTMR10,OTUD7A,TRPM1 | fam_ctrl | 2 | EG001 | 1 |
| chr15:30936285-32514341 | 1578057 | AK093758,ARHGAP11B,CHRNA7,DKFZp434L187,DQ572979,DQ582939,DQ588973,DQ595055,DQ596686,DQ600342,DQ786280,FAN1,HERC2P10,JB175342,KLF13,LOC100288637,LOC283710,MIR211,MTMR10,OTUD7A,TRPM1 | fam_ctrl | 2 | EG001 | 1 |
| chr15:30936285-32514341 | 1578057 | AK093758,ARHGAP11B,CHRNA7,DKFZp434L187,DQ572979,DQ582939,DQ588973,DQ595055,DQ596686,DQ600342,DQ786280,FAN1,HERC2P10,JB175342,KLF13,LOC100288637,LOC283710,MIR211,MTMR10,OTUD7A,TRPM1 | GGE | 1 | EG001 | 1 |
| chr15:30936285-32514341 | 1578057 | AK093758,ARHGAP11B,CHRNA7,DKFZp434L187,DQ572979,DQ582939,DQ588973,DQ595055,DQ596686,DQ600342,DQ786280,FAN1,HERC2P10,JB175342,KLF13,LOC100288637,LOC283710,MIR211,MTMR10,OTUD7A,TRPM1 | GGE | 2 | EG001 | 1 |
| chr15:30936285-32514341 | 1578057 | AK093758,ARHGAP11B,CHRNA7,DKFZp434L187,DQ572979,DQ582939,DQ588973,DQ595055,DQ596686,DQ600342,DQ786280,FAN1,HERC2P10,JB175342,KLF13,LOC100288637,LOC283710,MIR211,MTMR10,OTUD7A,TRPM1 | GGE | 2 | EG087 | 1 |
| chr15:30936285-32514341 | 1578057 | AK093758,ARHGAP11B,CHRNA7,DKFZp434L187,DQ572979,DQ582939,DQ588973,DQ595055,DQ596686,DQ600342,DQ786280,FAN1,HERC2P10,JB175342,KLF13,LOC100288637,LOC283710,MIR211,MTMR10,OTUD7A,TRPM1 | GGE | 1 | EG282 | 1 |
| chr15:30936285-32514341 | 1578057 | AK093758,ARHGAP11B,CHRNA7,DKFZp434L187,DQ572979,DQ582939,DQ588973,DQ595055,DQ596686,DQ600342,DQ786280,FAN1,HERC2P10,JB175342,KLF13,LOC100288637,LOC283710,MIR211,MTMR10,OTUD7A,TRPM1 | GGE | 2 | EG328 | 1 |
| chr15:30936285-32514341 | 1578057 | AK093758,ARHGAP11B,CHRNA7,DKFZp434L187,DQ572979,DQ582939,DQ588973,DQ595055,DQ596686,DQ600342,DQ786280,FAN1,HERC2P10,JB175342,KLF13,LOC100288637,LOC283710,MIR211,MTMR10,OTUD7A,TRPM1 | GGE | 1 | EG360 | 1 |
| chr15:30936285-32514341 | 1578057 | AK093758,ARHGAP11B,CHRNA7,DKFZp434L187,DQ572979,DQ582939,DQ588973,DQ595055,DQ596686,DQ600342,DQ786280,FAN1,HERC2P10,JB175342,KLF13,LOC100288637,LOC283710,MIR211,MTMR10,OTUD7A,TRPM1 | GGE | 1 | EG430 | 1 |
| chr15:32922947-32971934 | 48988 | NOT_FOUND | fam_ctrl | 1 | EP013 | 0 |
| chr16:15092778-15225383 | 132606 | FLJ00285,NPIP,NTAN1,PDXDC1,RRN3 | GGE | 2 | EG406 | 1 |
| chr16:15493046-16291983 | 798938 | ABCC1,ABCC6,AX747846,C16orf45,FOPNL,KIAA0430,MIR484,MPV17L,MYH11,NDE1 | GGE | 2 | EG406 | 1 |
| chr16:15493046-18164698 | 2671653 | ABCC1,ABCC6,AK310228,AX747757,AX747846,C16orf45,DQ586919,DQ596229,FOPNL,KIAA0430,MIR3179-2,MIR3180-3,MIR484,MPV17L,MYH11,Mir_548,NDE1,NOMO3,NPIP,PKD1P1,XYLT1 | fam_ctrl | 2 | EG108 | 1 |
| chr16:15493046-18164698 | 2671653 | ABCC1,ABCC6,AK310228,AX747757,AX747846,C16orf45,DQ586919,DQ596229,FOPNL,KIAA0430,MIR3179-2,MIR3180-3,MIR484,MPV17L,MYH11,Mir_548,NDE1,NOMO3,NPIP,PKD1P1,XYLT1 | fam_ctrl | 1 | EG108 | 1 |
| chr16:15493046-18164698 | 2671653 | ABCC1,ABCC6,AK310228,AX747757,AX747846,C16orf45,DQ586919,DQ596229,FOPNL,KIAA0430,MIR3179-2,MIR3180-3,MIR484,MPV17L,MYH11,Mir_548,NDE1,NOMO3,NPIP,PKD1P1,XYLT1 | GGE | 2 | EG108 | 1 |
| chr16:15493046-18164698 | 2671653 | ABCC1,ABCC6,AK310228,AX747757,AX747846,C16orf45,DQ586919,DQ596229,FOPNL,KIAA0430,MIR3179-2,MIR3180-3,MIR484,MPV17L,MYH11,Mir_548,NDE1,NOMO3,NPIP,PKD1P1,XYLT1 | GGE | 2 | EG108 | 1 |
| chr16:28825605-29042014 | 216410 | AK125489,ATP2A1,ATXN2L,CD19,LAT,LOC100289092,MIR4517,MIR4721,NFATC2IP,NPIPL1,RABEP2,SH2B1,SPNS1,TUFM | ctrl | 2 | NA | 1 |
| chr16:29652488-30192359 | 539872 | AB209061,AK097453,AK097472,AK097527,ALDOA,ASPHD1,BC029255,BC041466,BOLA2,C16orf54,C16orf92,CDIPT,CDIPT-AS1,DOC2A,FAM57B,GDPD3,HIRIP3,INO80E,KCTD13,KIF22,MAPK3,MAZ,MVP,PAGR1,PPP4C,PRRT2,QPRT,SEZ6L2,SPN,TAOK2,TBX6,TMEM219,YPEL3,ZG16 | Mixed | 2 | EG067 | 1 |
| chr17:33684035-33768199 | 84165 | SLFN11,SLFN12,SLFN13 | ctrl | 2 | NA | 1 |
| chr17:33684035-33768199 | 84165 | SLFN11,SLFN12,SLFN13 | ctrl | 1 | NA | 1 |
| chr17:33684035-33768199 | 84165 | SLFN11,SLFN12,SLFN13 | ctrl | 1 | NA | 1 |
| chr17:33684035-33768199 | 84165 | SLFN11,SLFN12,SLFN13 | ctrl | 1 | NA | 1 |
| chr17:33684035-33768199 | 84165 | SLFN11,SLFN12,SLFN13 | ctrl | 2 | NA | 1 |
| chr17:33684035-33768199 | 84165 | SLFN11,SLFN12,SLFN13 | ctrl | 2 | NA | 1 |
| chr17:33684035-33768199 | 84165 | SLFN11,SLFN12,SLFN13 | GGE | 2 | EG386 | 1 |
| chr17:33684035-33768199 | 84165 | SLFN11,SLFN12,SLFN13 | GGE | 1 | EG396 | 1 |
| chr17:33684035-33768199 | 84165 | SLFN11,SLFN12,SLFN13 | GGE | 2 | EG406 | 1 |
| chr17:33684035-33768199 | 84165 | SLFN11,SLFN12,SLFN13 | GGE | 1 | EG417 | 1 |
| chr17:33684035-33768199 | 84165 | SLFN11,SLFN12,SLFN13 | NAFE | 2 | EP300 | 1 |
| chr22:18115392-18633446 | 518055 | BC064400,BCL2L13,BID,DQ570096,Em:AC008101.5,FLJ41941,MICAL3,MIR3198-1,MIR648,PEX26,TUBA8,USP18 | GGE | 2 | EG292 | 1 |
| chr22:18889490-21463730 | 2574241 | 7SK,AIFM3,AK129567,AK302545,ARVCF,BC033281,BC035867,BC127858,BCRP2,BX648073,C22orf29,C22orf39,CDC45,CLDN5,CLTCL1,COMT,CRKL,DGCR10,DGCR11,DGCR14,DGCR2,DGCR5,DGCR6,DGCR6L,DGCR8,DGCR9,DQ571461,DQ574263,DQ585141,GNB1L,GSC2,<br>HIRA,HV593096,HV593110,HV593127,HV593134,HV593135,HV593178,HV593183,JX456220,KIAA1653,KLHL22,LINC00895,LINC00896,LOC284865,LOC388849,LOC400891,LOC729444,LZTR1,MED15,MIR1286,MIR1306,MIR185,MIR3618,MIR4761,MRPL40,Metazoa_SRP,<br>Mir_649,P2RX6,P2RX6P,PI4KA,PI4KAP1,POM121L4P,PRODH,RANBP1,RIMBP3,RTN4R,SCARF2,SEPT5-GP1BB,SERPIND1,SLC25A1,SLC7A4,SNAP29,TANGO2,TBX1,THAP7,THAP7-AS1,TMEM191A,TMEM191B,TRMT2A,TSSK2,TUBA3FP,TXNRD2,U84523,UFD1L,USP41,Y_RNA,ZDHHC8,ZNF74 | Mixed | 1 | EP016 | 1 |
| chr22:22314463-22362353 | 47891 | AK131325,TOP3B | ctrl | 2 | NA | 1 |
| chr22:22314463-22379067 | 64605 | AK131325,TOP3B | ctrl | 1 | NA | 1 |
| chr22:22314463-22379067 | 64605 | AK131325,TOP3B | ctrl | 1 | NA | 1 |
| chr22:22314463-22379067 | 64605 | AK131325,TOP3B | ctrl | 1 | NA | 1 |
| chr22:22314463-22379067 | 64605 | AK131325,TOP3B | ctrl | 1 | NA | 1 |
| chr22:22314463-22379067 | 64605 | AK131325,TOP3B | GGE | 2 | EG072 | 1 |
| chr22:22314463-22379067 | 64605 | AK131325,TOP3B | Mixed | 1 | EP244 | 1 |

Orange = Same individual
