## Supplemental Table 4 for "Assessment of burden and segregation profiles of CNVs in patients with epilepsy"

| CNV | Length | Genes | Phenotype | Sex | Family | Coding | In intolerant gene | In epilepsy gene | Hotspot | WGS | WES |
|---|---|---|---|---|---|---|---|---|---|---|---|
| chr10:129688467-130926027 | 1237561 | AK124226,CLRN3,MKI67,PTPRE | GGE | 1 | EG058 | 1 | 0 | 0 | 0 | 0 | 0 |
| chr10:129688467-130926027 | 1237561 | AK124226,CLRN3,MKI67,PTPRE | GGE | 2 | EG058 | 1 | 0 | 0 | 0 | 0 | 0 |
| chr13:23515553-24927971 | 1412419 | ANKRD20A19P,BC038727,BC043582,C1QTNF9,C1QTNF9B,C1QTNF9B-AS1,LINC00327,MIPEP,MIR2276,SACS,SACS-AS1,SGCG,SPATA13,SPATA13-AS1,TNFRSF19 | fam_ctrl | 2 | EG012 | 1 | 0 | 0 | 0 | 0 | 0 |
| chr13:23526039-24927971 | 1401933 | ANKRD20A19P,BC038727,BC043582,C1QTNF9,C1QTNF9B,C1QTNF9B-AS1,LINC00327,MIPEP,MIR2276,SACS,SACS-AS1,SGCG,SPATA13,SPATA13-AS1,TNFRSF19 | GGE | 2 | EG012 | 1 | 0 | 0 | 0 | 0 | 0 |
| chr13:23526039-24927971 | 1401933 | ANKRD20A19P,BC038727,BC043582,C1QTNF9,C1QTNF9B,C1QTNF9B-AS1,LINC00327,MIPEP,MIR2276,SACS,SACS-AS1,SGCG,SPATA13,SPATA13-AS1,TNFRSF19 | GGE | 2 | EG012 | 1 | 0 | 0 | 0 | 1 | 0 |
| chr15:30936285-32514341 | 1578057 | AK093758,ARHGAP11B,CHRNA7,DKFZp434L187,DQ572979,DQ582939,DQ588973,DQ595055,DQ596686,DQ600342,DQ786280,FAN1,HERC2P10,JB175342,KLF13,LOC100288637,LOC283710,MIR211,MTMR10,OTUD7A,TRPM1 | fam_ctrl | 2 | EG001 | 1 | 0 | 1 | 15q13.3 | 0 | 0 |
| chr15:30936285-32514341 | 1578057 | AK093758,ARHGAP11B,CHRNA7,DKFZp434L187,DQ572979,DQ582939,DQ588973,DQ595055,DQ596686,DQ600342,DQ786280,FAN1,HERC2P10,JB175342,KLF13,LOC100288637,LOC283710,MIR211,MTMR10,OTUD7A,TRPM1 | fam_ctrl | 2 | EG001 | 1 | 0 | 1 | 15q13.3 | 0 | 0 |
| chr15:30936285-32514341 | 1578057 | AK093758,ARHGAP11B,CHRNA7,DKFZp434L187,DQ572979,DQ582939,DQ588973,DQ595055,DQ596686,DQ600342,DQ786280,FAN1,HERC2P10,JB175342,KLF13,LOC100288637,LOC283710,MIR211,MTMR10,OTUD7A,TRPM1 | GGE | 1 | EG001 | 1 | 0 | 1 | 15q13.3 | 0 | 0 |
| chr15:30936285-32514341 | 1578057 | AK093758,ARHGAP11B,CHRNA7,DKFZp434L187,DQ572979,DQ582939,DQ588973,DQ595055,DQ596686,DQ600342,DQ786280,FAN1,HERC2P10,JB175342,KLF13,LOC100288637,LOC283710,MIR211,MTMR10,OTUD7A,TRPM1 | GGE | 2 | EG001 | 1 | 0 | 1 | 15q13.3 | 1 | 0 |
| chr15:30936285-32514341 | 1578057 | AK093758,ARHGAP11B,CHRNA7,DKFZp434L187,DQ572979,DQ582939,DQ588973,DQ595055,DQ596686,DQ600342,DQ786280,FAN1,HERC2P10,JB175342,KLF13,LOC100288637,LOC283710,MIR211,MTMR10,OTUD7A,TRPM1 | GGE | 2 | EG087 | 1 | 0 | 1 | 15q13.3 | 0 | 1 |
| chr15:30936285-32514341 | 1578057 | AK093758,ARHGAP11B,CHRNA7,DKFZp434L187,DQ572979,DQ582939,DQ588973,DQ595055,DQ596686,DQ600342,DQ786280,FAN1,HERC2P10,JB175342,KLF13,LOC100288637,LOC283710,MIR211,MTMR10,OTUD7A,TRPM1 | GGE | 1 | EG282 | 1 | 0 | 1 | 15q13.3 | 0 | 1 |
| chr15:30936285-32514341 | 1578057 | AK093758,ARHGAP11B,CHRNA7,DKFZp434L187,DQ572979,DQ582939,DQ588973,DQ595055,DQ596686,DQ600342,DQ786280,FAN1,HERC2P10,JB175342,KLF13,LOC100288637,LOC283710,MIR211,MTMR10,OTUD7A,TRPM1 | GGE | 1 | EG360 | 1 | 0 | 1 | 15q13.3 | 0 | 1 |
| chr15:30936285-32514341 | 1578057 | AK093758,ARHGAP11B,CHRNA7,DKFZp434L187,DQ572979,DQ582939,DQ588973,DQ595055,DQ596686,DQ600342,DQ786280,FAN1,HERC2P10,JB175342,KLF13,LOC100288637,LOC283710,MIR211,MTMR10,OTUD7A,TRPM1 | GGE | 2 | EG328 | 1 | 0 | 1 | 15q13.3 | 1 | 0 |
| chr15:30936285-32514341 | 1578057 | AK093758,ARHGAP11B,CHRNA7,DKFZp434L187,DQ572979,DQ582939,DQ588973,DQ595055,DQ596686,DQ600342,DQ786280,FAN1,HERC2P10,JB175342,KLF13,LOC100288637,LOC283710,MIR211,MTMR10,OTUD7A,TRPM1 | GGE | 1 | EG430 | 1 | 0 | 1 | 15q13.3 | 1 | 0 |
| chr16:15493046-18164698 | 2671653 | ABCC1,ABCC6,AK310228,AX747757,AX747846,C16orf45,DQ586919,DQ596229,FOPNL,KIAA0430,MIR3179-2,MIR3180-3,MIR484,MPV17L,MYH11,Mir_548,NDE1,NOMO3,NPIP,PKD1P1,XYLT1 | fam_ctrl | 1 | EG108 | 1 | 1 | 0 | 16p13.11 | 0 | 0 |
| chr16:15493046-18164698 | 2671653 | ABCC1,ABCC6,AK310228,AX747757,AX747846,C16orf45,DQ586919,DQ596229,FOPNL,KIAA0430,MIR3179-2,MIR3180-3,MIR484,MPV17L,MYH11,Mir_548,NDE1,NOMO3,NPIP,PKD1P1,XYLT1 | fam_ctrl | 2 | EG108 | 1 | 1 | 0 | 16p13.11 | 0 | 0 |
| chr16:15493046-18164698 | 2671653 | ABCC1,ABCC6,AK310228,AX747757,AX747846,C16orf45,DQ586919,DQ596229,FOPNL,KIAA0430,MIR3179-2,MIR3180-3,MIR484,MPV17L,MYH11,Mir_548,NDE1,NOMO3,NPIP,PKD1P1,XYLT1 | GGE | 2 | EG108 | 1 | 1 | 0 | 16p13.11 | 0 | 0 |
| chr16:15493046-18164698 | 2671653 | ABCC1,ABCC6,AK310228,AX747757,AX747846,C16orf45,DQ586919,DQ596229,FOPNL,KIAA0430,MIR3179-2,MIR3180-3,MIR484,MPV17L,MYH11,Mir_548,NDE1,NOMO3,NPIP,PKD1P1,XYLT1 | GGE | 2 | EG108 | 1 | 1 | 0 | 16p13.11 | 1 | 0 |
| chr17:14101029-15471179 | 1370151 | BC037357,CDRT15,CDRT4,CDRT7,COX10,HS3ST3B1,MGC12916,MIR4731,Metazoa_SRP,PMP22,TEKT3,TRNA_Sup,TVP23C,TVP23C-CDRT4 | Mixed | 1 | EG389 | 1 | 0 | 0 | 0 | 1 | 0 |
| chr18:64525217-77553173 | 13027957 | AK094611,ATP9B,AX747775,BC037384,BC047599,BC070490,C18orf63,CBLN2,CCDC102B,CD226,CNDP1,CNDP2,CTDP1,CYB5A,DOK6,DSEL,FAM69C,FBXO15,FLJ44313,FLJ44881,GALR1,LINC00908,LINC00909,LOC100131655,LOC100505776,LOC100505817,LOC339298,LOC400655,LOC643542,MBP,MIR5011,Metazoa_SRP,NETO1,NFATC1,RTTN,SALL3,SMIM21,SOCS6,TIMM21,TMX3,TSHZ1,ZADH2,ZNF236,ZNF407,ZNF516 | NAFE | 2 | EP014 | 1 | 1 | 0 | 0 | 0 | 1 |
| chr2:194098094-195484228 | 1386135 | BC038548 | ctrl | 2 | NA | 1 | 0 | 0 | 0 | 0 | 0 |
| chr22:18889490-21463730 | 2574241 | 7SK,AIFM3,AK129567,AK302545,ARVCF,BC033281,BC035867,BC127858,BCRP2,BX648073,C22orf29,C22orf39,CDC45,CLDN5,CLTCL1,COMT,CRKL,DGCR10,DGCR11,DGCR14,DGCR2,DGCR5,DGCR6,DGCR6L,DGCR8,DGCR9,DQ571461,DQ574263,DQ585141,GNB1L,GSC2,HIRA,HV593096,HV593110,HV593127,HV593134,HV593135,HV593178,HV593183,JX456220,KIAA1653,KLHL22,LINC00895,LINC00896<br>LOC284865,LOC388849,LOC400891,LOC729444,LZTR1,MED15,MIR1286,MIR1306,MIR185,MIR3618,MIR4761,MRPL40,Metazoa_SRP,Mir_649,P2RX6,P2RX6P,PI4KA,PI4KAP1,POM121L4P,PRODH,RANBP1,RIMBP3,RTN4R,SCARF2,SEPT5-GP1BB,SERPIND1,SLC25A1,SLC7A4,SNAP29,TANGO2,TBX1,THAP7,THAP7-AS1,TMEM191A,TMEM191B,TRMT2A,TSSK2,TUBA3FP,TXNRD2,U84523,UFD1L,USP41,Y_RNA,ZDHHC8,ZNF74 | Mixed | 1 | EP016 | 1 | 1 | 0 | 22q11.2 | 0 | 0 |
| chr3:156777225-158554978 | 1777754 | 7SK,AK097794,AY070437,C3orf55,CCNL1,GFM1,LINC00880,LINC00881,LXN,MFSD1,MLF1,Mir_584,PTX3,RARRES1,RSRC1,SHOX2,VEPH1 | GGE | 1 | EG121 | 1 | 0 | 0 | 0 | 0 | 1 |
| chr3:78322422-79522372 | 1199951 | NOT_FOUND | ctrl | 1 | NA | 0 | 0 | 0 | 0 | 0 | 0 |
| chr7:108854537-109969407 | 1114871 | EIF3IP1 | NAFE | 1 | EP062 | 1 | 0 | 0 | 0 | 0 | 0 |
| chr7:108854537-109984004 | 1129468 | EIF3IP1 | Mixed | 1 | EP062 | 1 | 0 | 0 | 0 | 0 | 0 |
| chr8:135644369-136843565 | 1199197 | KHDRBS3,LOC286094,MIR30B,MIR30D,Mir_652,U1,ZFAT | DEE_trio_parent | 1 | Trio88 | 1 | 0 | 0 | 0 | 1 | 0 |
| chr9:31111743-32128828 | 1017086 | NOT_FOUND | ctrl | 2 | NA | 0 | 0 | 0 | 0 | 0 | 0 |
