## Supplemental Table 5 for "Assessment of burden and segregation profiles of CNVs in patients with epilepsy"

| Chromosome | Position | Aleternate allele | Phenotype | WGS/WES | CNET cohort frequency | gnomAD frequency | gnomAD clinical significance | UNEECON score |
|---|---|---|---|---|---|---|---|---|
| 17 | 15234895 | T | Mixed | WGS | 0.25 | 0.21 | NA | 0.000081 |
| 18 | 74980809 | A | NAFE | WES | 1.00 | 0.97 | NA | 0.000109 |
| 18 | 72228124 | G | NAFE | WES | 1.00 | 0.99 | NA | 0.000111 |
| 13 | 24895559 | G | GGE | WGS | 0.74 | 0.76 | NA | 0.000442 |
| 16 | 16271357 | C | GGE | WGS | 1.00 | 0.99 | benign | 0.000522 |
| 3 | 158388780 | C | GGE | WES | 0.53 | 0.52 | NA | 0.000650 |
| 13 | 23898664 | G | GGE | WGS | 0.87 | 0.88 | benign | 0.000700 |
| 18 | 72114455 | T | NAFE | WES | 1.00 | 0.99 | NA | 0.000802 |
| 15 | 31362352 | T | GGE | WES | 0.81 | 0.81 | benign | 0.000931 |
| 15 | 31362352 | T | GGE | WGS | 0.80 | 0.81 | benign | 0.000931 |
| 15 | 31362352 | T | GGE | WES | 0.81 | 0.81 | benign | 0.000931 |
| 15 | 31362352 | T | GGE | WES | 0.81 | 0.81 | benign | 0.000931 |
| 15 | 31362352 | T | GGE | WGS | 0.80 | 0.81 | benign | 0.000931 |
| 15 | 31362352 | T | GGE | WGS | 0.80 | 0.81 | benign | 0.000931 |
| 18 | 74611127 | G | NAFE | WES | 0.96 | 0.95 | NA | 0.001479 |
| 3 | 158366900 | A | GGE | WES | 0.53 | 0.55 | benign | 0.001635 |
| 18 | 66504459 | T | NAFE | WES | 1.00 | 0.98 | NA | 0.001985 |
| 18 | 72103782 | C | NAFE | WES | 0.38 | 0.35 | NA | 0.002147 |
| 18 | 72201918 | A | NAFE | WES | 0.12 | 0.11 | NA | 0.004205 |
| 17 | 14139891 | C | Mixed | WGS | 1.00 | 0.98 | NA | 0.004650 |
| 18 | 72021717 | C | NAFE | WES | 1.00 | 1.00 | NA | 0.005075 |
| 13 | 24411772 | C | GGE | WGS | 0.99 | 0.99 | NA | 0.007212 |
| 18 | 76753768 | G | NAFE | WES | 0.82 | 0.84 | NA | 0.012774 |
| 18 | 66513615 | G | NAFE | WES | 0.38 | 0.38 | NA | 0.015114 |
| 15 | 31197564 | A | GGE | WES | 0.44 | 0.46 | benign | 0.017849 |
| 15 | 31197564 | A | GGE | WES | 0.44 | 0.46 | benign | 0.017849 |
| 15 | 31197564 | A | GGE | WGS | 0.43 | 0.46 | benign | 0.017849 |
| 18 | 67718688 | G | NAFE | WES | 0.95 | 0.91 | benign | 0.020459 |
| 18 | 72998899 | A | NAFE | WES | 0.03 | 0.03 | benign | 0.023956 |
| 18 | 70417396 | T | NAFE | WES | 1.00 | 1.00 | NA | 0.026219 |
| 18 | 77473127 | T | NAFE | WES | 0.18 | 0.14 | benign | 0.031721 |
| 3 | 157160196 | G | GGE | WES | 7.4e-04 | 1.6e-05 | NA | 0.047242 |
| 18 | 67871343 | C | NAFE | WES | 0.92 | 0.86 | benign | 0.064903 |
| 18 | 76753588 | G | NAFE | WES | 0.84 | 0.79 | benign | 0.107196 |
| 15 | 31776021 | C | GGE | WGS | 0.99 | 0.99 | NA | 0.107602 |
| 15 | 31776021 | C | GGE | WGS | 0.99 | 0.99 | NA | 0.107602 |
| 15 | 31776021 | C | GGE | WGS | 0.99 | 0.99 | NA | 0.107602 |
| 13 | 24798506 | T | GGE | WGS | 0.03 | 0.02 | NA | 0.119246 |
| 18 | 77246406 | G | NAFE | WES | 0.49 | 0.42 | NA | 0.142239 |
| 17 | 15142755 | A | Mixed | WGS | 0.09 | 0.08 | benign | #N/A |
| 17 | 15134175 | G | Mixed | WGS | 0.53 | 0.53 | benign | #N/A |
